# Demonstration of a portable intracortical brain-computer interface

**DOI:** 10.1101/19004721

**Authors:** Jeffrey M. Weiss, Robert A. Gaunt, Robert Franklin, Michael Boninger, Jennifer L. Collinger

## Abstract

While recent advances in intracortical brain-computer interfaces (iBCI) have demonstrated the ability to restore motor and communication functions, such demonstrations have generally been confined to controlled experimental settings and have required bulky laboratory hardware. Here, we developed and evaluated a self-contained portable iBCI that enabled the user to interact with various computer programs. The iBCI, which weighs 1.5 kg, consists of digital headstages, a small signal processing hub, and a tablet PC. A human participant tested the portable iBCI in laboratory and home settings under an FDA Investigational Device Exemption (NCT01894802). The participant successfully completed 96% of trials in a 2D cursor center-out task with the portable iBCI, a rate indistinguishable from that achieved with the standard laboratory iBCI. The participant also completed a variety of free-form tasks, including drawing, gaming, and typing.

## Introduction

Intracortical brain-computer interfaces (iBCI) are a promising technology to restore movement and communication abilities to individuals with significant neurological injuries or disease. Additionally, iBCI has restored movement to individuals with upper limb paralysis by using M1 signals to control functional electrical stimulation systems [1], [2] or robotic limbs [3], [4] with as many as 10 continuously controlled degrees-of-freedom [5]. However, all of these demonstrations depended on experienced technicians to operate large and complicated systems comprised of multiple computers, displays, signal processors, amplifiers, headstages, power supplies, and cables. The size and complexity of such a system is a major barrier that must be addressed before translating this technology into a take-home system.

The lack of a portable and convenient iBCI that is capable of functioning in many environments has been previously identified as a barrier to translation [6]. Additionally, potential iBCI users with upper limb paralysis have identified independent operation [7] and a simple setup [8] as important design characteristics of a final system. Wireless and/or portable BCIs, particularly in the non-invasive electroencephalographic (EEG) domain, have been developed [9]– [13], but few have been tested outside of the laboratory. Several case studies have sent users home with EEG [14], [15] or electrocorticographic (ECoG) [16] BCIs that could be operated on a daily basis without experimenters or technicians present. Intracortical BCIs with external wireless components have been demonstrated [17], [18], but still require large laboratory systems for data acquisition and processing and hence are not fully portable. Wireless implantable iBCIs that communicate with external processing hardware have also been developed [19], but so far have not been tested in humans. The development of an iBCI that is sufficiently portable and simple to set up and use is a required prerequisite to a long-term take-home study of iBCI use.

Here, we created and demonstrated a compact and portable iBCI that builds on FDA-cleared technology (Blackrock NeuroPort Biopotential Signal Processing System, Blackrock Microsystems, Salt Lake City, UT) used in human clinical trials. Our system serves as a proof-of-concept of a completely mobile system that could be used by individuals with locked-in-syndrome or upper limb paralysis for communication, computer use and recreation, and controlling external devices.

## Methods

### System components

The portable iBCI (Figure 1a) consists of digital headstages to acquire neural signals, a tablet PC, and a signal processor hub, which provides a data interface and power isolation between the headstages and tablet. Two Cereplex-E digital headstages (Blackrock Microsystems) were used to acquire neural signals from up to 256 electrodes. The wearable neural signal processor hub (wNSP hub) connects to each headstage via an HDMI cable to provide power and timing signals to the headstages and receive data from each electrode. The wNSP hub receives power and transmits data to the tablet PC via USB 2.0. The medical-grade tablet PC (Getac RX10H, Irvine, CA) is IEC 60601-1 compliant and runs Windows 10. The tablet’s battery powers the entire system. The hardware components weigh a total of 1.5 kg. All digital signal processing, decoding, and user interaction occur on the tablet. As a result, this system is completely portable and does not require any remote connection to other computer systems. Figure 1 compares the portable iBCI components with our full laboratory system, which consists of a large rack of PCs and neural signal processors, multiple displays, analog patient cable headstages, and a rack containing large amplifiers and power supplies.

**Figure 1.**
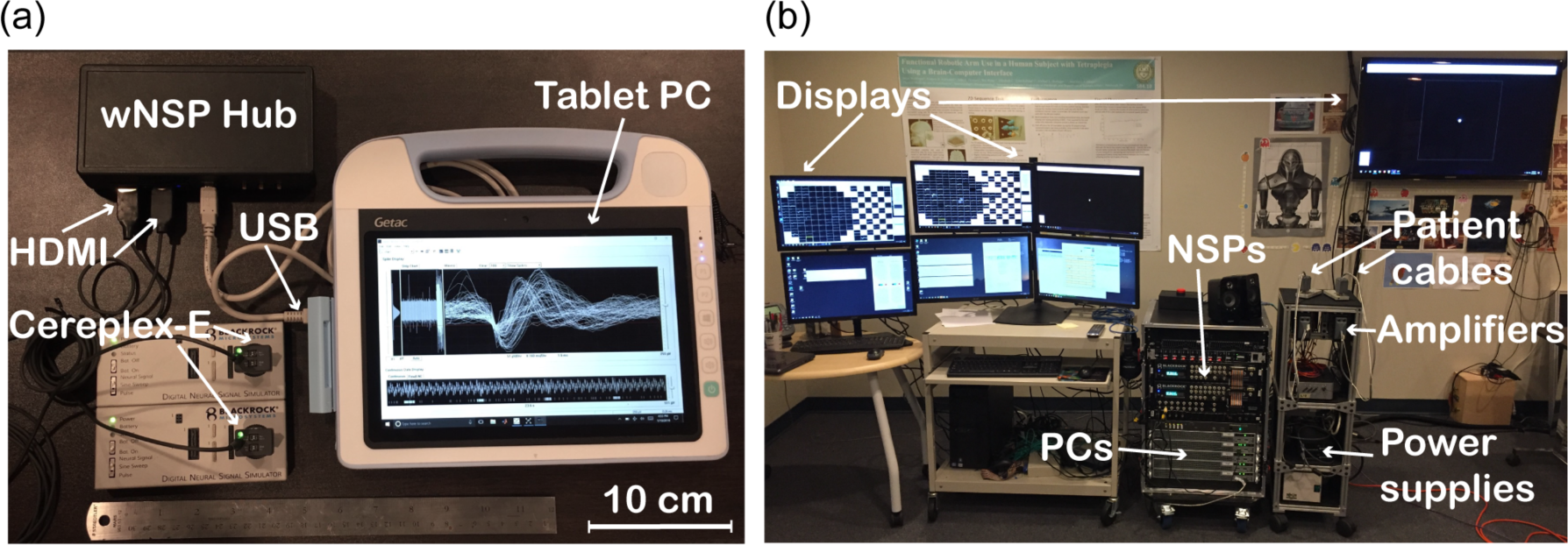
Components of portable and standard iBCI systems. (a) The portable iBCI consists of a tablet PC, wNSP hub, and two Cereplex-E headstages. The headstages are connected to two neural signal simulators in place of an iBCI user. The headstages are connected to the wNSP hub via HDMI cables, and the wNSP hub is connected to the tablet via USB. A 12-inch ruler is shown for reference. (b) Our standard iBCI is much larger, and consists of two neural signal processors (NSPs), patient cable headstages, amplifiers, power supplies, and multiple PCs and displays. Note that the photograph of our standard iBCI also includes stimulation hardware (i.e., a stimulator and PC) not replicated in the portable iBCI.

### Software overview

The tablet PC runs the Blackrock Central Suite and nPlay Server (Blackrock Microsystems) to acquire, filter, and detect spikes from each recording channel. nPlay Server acquires real-time neural voltage data over USB from up to 256 channels continuously sampled at 30 kHz. We configured the system via the Central Suite to filter the voltage data with a first-order Butterworth 750 Hz high-pass filter and apply a spike detection threshold at −4.5 times the root-mean-square voltage [20], estimated during initial setup from two seconds of filtered data using an algorithm developed by Blackrock Microsystems [21]. The system can also be configured to perform online spike sorting, but the sorting feature was not tested in this study since our decoding scheme uses threshold crossings on a per channel basis. Extracted spike snippets are accessible from other applications via a software development kit (cbSDK, Blackrock Microsystems).

A modular iBCI software suite, adapted from a system described previously [3], received and decoded spike data from nPlay Server to control a cursor, mouse emulator, or keypress emulator. The iBCI software suite, which is summarized in Figure 2, consists of several modules programmed in C++ and Matlab (MathWorks, Natick, MA). All modules communicate with each other using the Dragonfly messaging architecture [22]. The iBCI suite acquires neural spike data from nPlay Server via cbSDK. Spike Processing Module 1 reads spike snippets from cbSDK and generates spike counts binned in 10 ms increments, a duration defined by a “heartbeat” packet received from cbSDK. Spike Processing Module 2 converts the bin width to 20 ms, a duration we defined to yield a 50 Hz system update rate, by summing pairs of spike count messages. The Decoder Module transforms neural firing rates into velocity control signals that can drive Mouse and Keypress Emulator Modules. This allows the iBCI user to interface with third-party Windows applications such as painting applications, computer games, or on-screen keyboards. The Decoder Module is also capable of displaying a center-out cursor environment and auto-generating velocity commands during the decoder calibration procedure. The overall system configuration and task progression is managed by the Task Executive Module.

**Figure 2.**
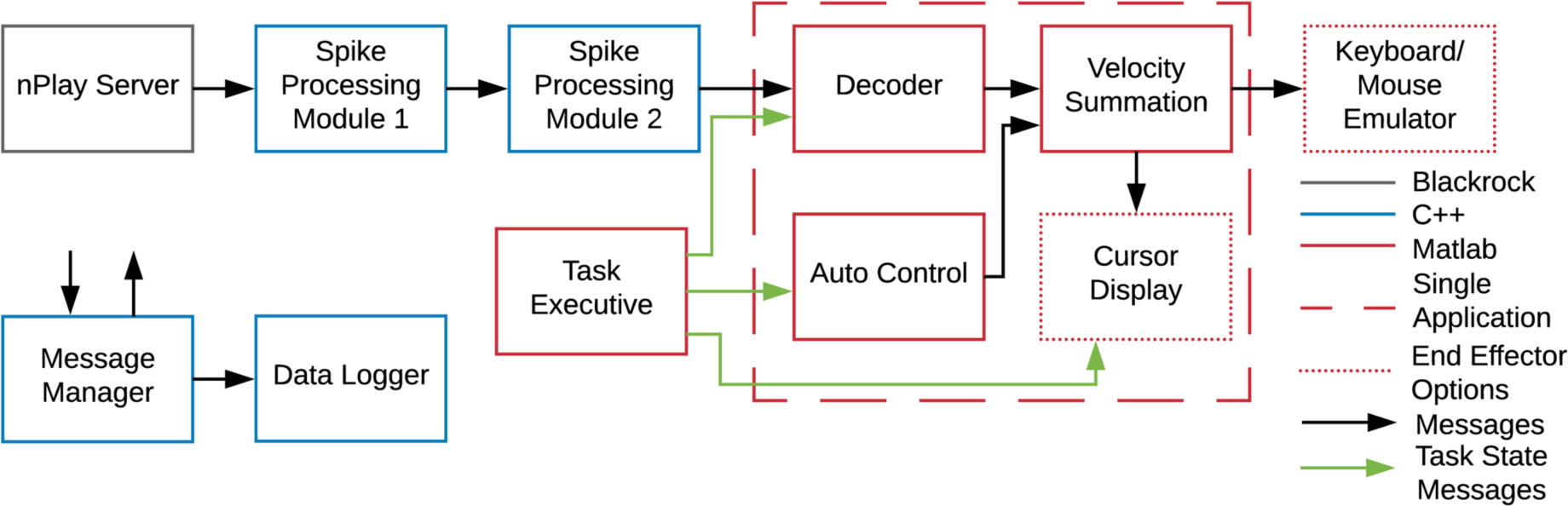
Portable iBCI software. Spike data is output from nPlay Server to Spike Processing Module 1 via cbSDK. All downstream messages were sent via the Dragonfly Messaging system, which uses Message Manager to coordinate messages between multiple modules. All messages are received by the Data Logger and saved for offline analysis. Some of the modules used in the standard iBCI were consolidated into a single application on the portable iBCI (dashed red box). All custom modules were written in C++ (blue) or Matlab (red). The “Task Executive” is responsible for controlling the state of the system and configuring other modules. Spike data is binned in the Spike Processing Modules and then decoded to generate a velocity command every 20 ms, which can drive a cursor, emulated mouse, or emulated keypresses.

### Human participant and implant

The portable iBCI was tested by a human participant in a study conducted under an Investigational Device Exemption granted by the U.S. Food and Drug Administration. The study received approval from the Institutional Review Boards at the University of Pittsburgh (Pittsburgh, PA) and the Space and Naval Warfare Systems Center Pacific (San Diego, CA) and is registered at ClinicalTrials.gov (NCT01894802). Informed consent was obtained prior to performing any experimental procedures. The participant consented to the use of still images or videos in publications related to this work.

A 28-year old male with a chronic C5 motor/C6 sensory ASIA B spinal cord injury was implanted with four intracortical microelectrode arrays (Blackrock Microsystems) in primary motor (M1) and primary somatosensory (S1) cortex, as described previously [23]. The arrays were wired to two pedestals in two pairs, each consisting of an 88-channel array in M1 and a 32-channel array in S1. A total of 240 channels were recorded across the four arrays, with 176 channels in M1 used for to decode intended movement signals. The participant began using the portable iBCI approximately three years after implant and was experienced in using the standard laboratory iBCI system. The portable iBCI was tested both in the lab and in the participant’s home with experimenters present to setup and supervise use of the system. All data presented in this manuscript were collected between 1058 and 1435 days post-implant.

### Signal quality comparison

The signal quality of the digital headstages and portable iBCI was compared against the standard laboratory system by quantifying the signal-to-noise ratio (SNR) of sorted units recorded on three system configurations: the laboratory system with analog patient cable headstages (standard system), the laboratory system with digital Cereplex-E headstages and digital hubs (digital system), and the portable iBCI system (portable system). Thirty seconds of resting baseline data were acquired in three experiment sessions for each system configuration. Spikes were extracted offline from continuous 30 kHz voltage recordings by filtering each signal with a fourth-order Butterworth 250 Hz high-pass filter and applying a threshold at −4.5 times the root-mean-square voltage, estimated from two seconds of filtered data using the same algorithm implemented in the Blackrock Central Suite [21]. Spikes were sorted offline using principal component analysis and a Gaussian mixture model expectation-maximization algorithm [24]. The SNR was calculated for each unit by dividing the mean peak-to-peak voltage (*V*_pp_) by two times the standard deviation of the baseline noise (σ_noise_), as measured from the filtered signal after removing all spike snippets. Units were sorted separately for each day and assumed to be independent. Within a day, units that were not recorded on all three recording systems were removed from further analysis. The SNR, *V*_pp_, and σ_noise_ for each unit were compared across system configurations using a one-way repeated measures analysis-of-variance (ANOVA). Post-hoc paired t-tests were completed to evaluate differences between recording modalities. We chose to use parametric tests even when data distributions were skewed because ANOVA is generally considered to be robust to non-normality and inequality of variance when sample sizes are large and balanced as is the case in our dataset [25]. Statistical testing was performed in R. All other data processing and analysis was performed in Matlab.

### System timing

System timing and delays were evaluated by analyzing timestamps from data packets recorded during a center-out cursor control task on the portable and standard systems. The output of Spike Processing Module 2, which consists of binned spike counts and is expected to be sent in regular 20 ms intervals based on the timing of “heartbeat” packets received from nPlay Server. The difference between the packet timestamps for sequential spike count messages was considered the system intersample period. The additional iBCI system processing and decoding delays were evaluated by calculating the difference between the binned spike count packet timestamps and subsequent decoded velocity command packet timestamps. This delay between the iBCI input and output was considered the system processing delay (SPD). The intersample period and SPD were compared between the standard system and portable iBCI tested in both the lab and at the participant’s home using a one-way ANOVA with post-hoc Tukey tests.

### Decoder training and center-out task

The iBCI was tested with an optimal linear estimator decoder to control a cursor with three degrees of freedom: 2D translation and click. A two-phase calibration procedure [3] was used in which the participant first observed, and then performed with error-assistance, a center-out task requiring the cursor to move from the center of the screen to one of eight targets arranged around a circle and presented in random order. Once the cursor successfully moved to each target, a random click target was presented by an audio cue (‘click’ or ‘unclick’). The click state of the cursor was indicated by a green circle which expanded around the cursor when clicked, and contracted to a point within the cursor when unclicked. During this task, click was represented as a continuously-varying kinematic variable, rather than a binary state. Decoders were trained after forty trials of observation, and again after forty trials of closed-loop error-assisted control, to relate each channel’s firing rate, *f*, estimated by 20 ms bins of threshold crossing counts, to velocity, *v*, using Equation (1). Firing rates were normalized by a square-root transform. Indirect optimal linear estimation with ridge regression was used to determine decoding weights, *b*, and the resulting decoder predicted velocity for *x* and *y* translation as well as click (*c*).

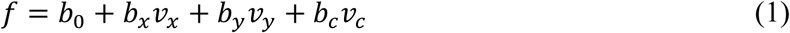

The decoder was first evaluated under brain-control using a 2D center-out task, which was identical to the training task, except that no click targets were presented and control of the click dimension was not evaluated. During each trial, the participant attempted to move the cursor to the randomly-presented target within 5 seconds. Forty trials of this task were performed during each session. The portable iBCI was used for three sessions at the lab and another three sessions at the participant’s home. An additional three sessions were conducted at the lab using the standard laboratory iBCI for comparison. For each successful trial, the time-to-target and normalized path length were calculated. Path lengths were normalized by the ideal path length, which was the minimum distance from the center of the circle to the edge of each target. The mean decoded normalized speed was calculated for all trials, including failed trials. Since the portable and laboratory systems have different monitor sizes and thus visually different target spacing, decoded speeds were normalized by the target distance (d). With this approach a constant normalized speed of 1 d/s along the ideal path would result in a time-to-target of 1 second. The decoded speed was equal to actual speed only if the intersample period was exactly 20 ms. Hence, the actual time-to-target was affected by three parameters: decoded speed, path length, and intersample period. The time-to-target, normalized path length, and normalized mean decoded speed metrics were compared across the three systems (standard laboratory iBCI, portable iBCI, and portable iBCI in the participant’s home) using one-way ANOVA tests with post-hoc Tukey tests.

### Functional evaluation

The 2D translation and click decoder trained on the portable iBCI was used to control mouse and keyboard emulators, which allowed the participant to perform freeform functional tasks of his choosing, including painting, typing with an on-screen keyboard, or computer games. The mouse emulator converted the decoded velocity commands to the position and click states of the Windows 10 system mouse cursor. The continuously-decoded click position, bounded between 0 and 1, was discretized into a clicked (position > 0.5) or unclicked (position < 0.5) state. The emulated mouse was used to control a painting application (TuxPaint) or to type using an on-screen keyboard. The keyboard emulator was used to control computer games by mapping positive and negative velocity commands for each degree of freedom to different emulated keypresses. A velocity threshold was applied to each direction (+*x*, −*x*, +*y*, −*y*, +*c*, −*c*) to trigger a unique keypress (e.g. arrow keys and spacebar) when the velocity crossed the threshold. The emulated keypresses were used as input to a variety of computer games of the participant’s choosing. Performance was not evaluated quantitatively, but sample performance was captured in the form of completed paintings and video capture of iBCI use.

## Results

### Signal quality

The SNR, peak-to-peak voltage (*V*_pp_), and baseline noise (σ_noise_) for each recording setup are quantified in Table 1. We found that recording setup had significant effects on both *V*_pp_ (F(2, 2704) = 236.4, *p* < 0.001) and σ_noise_ (F(2, 1438) = 58.16, *p* < 0.001), with significant differences between all system pairs (all *p* < 0.001). The changes in *V*_pp_ and σ_noise_ both resulted in a change in SNR. The baseline noise decreased with the digital headstages, and was lowest with the portable system, which had a 20% decrease in σ_noise_ compared to the standard system. However, the peak-to-peak voltage also decreased with the digital headstages such that the portable system had an 11% decrease in *V*_pp_ compared to the standard system. Overall, these changes yielded a 5% increase in SNR for the portable iBCI as compared to the standard system (F(2, 2704) = 128.4, *p* < 0.001; all post-hoc *p* < 0.001). As shown in Figure 3, the digital headstages systematically increased the SNR of sorted units for both the portable and laboratory systems.

**Table 1.**
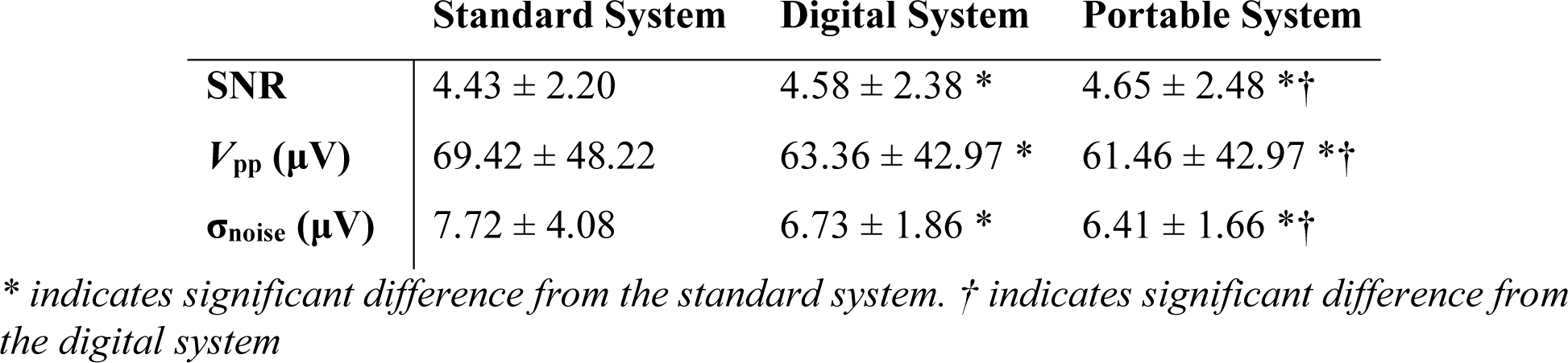
Signal quality metrics (mean and standard deviation)

**Figure 3.**
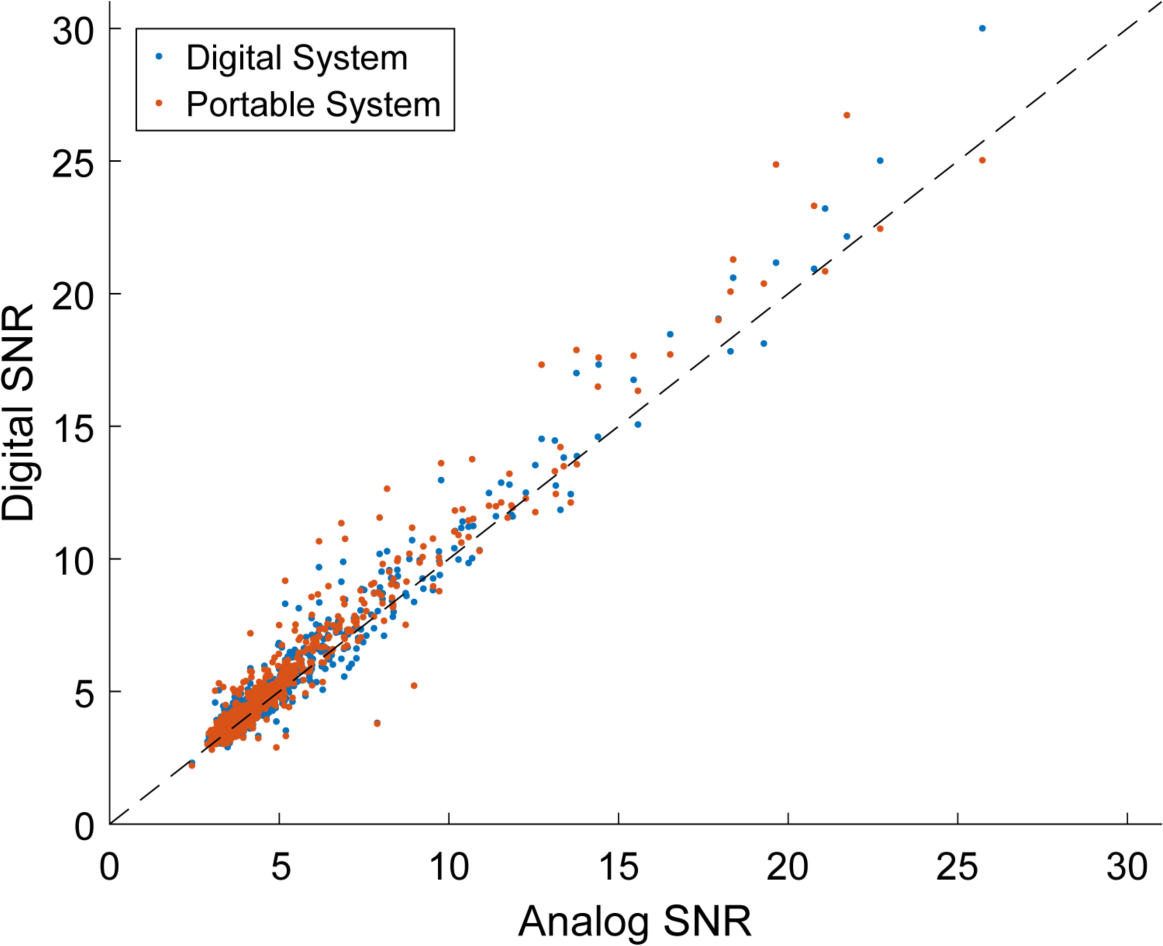
Signal-to-noise ratio of each unit recorded using the Cereplex-E digital headstage with the digital system (blue dots) and portable system (orange dots) plotted against the SNR recorded using analog patient cables and the standard system. The dashed line indicates the line where Digital SNR = Analog SNR. For most units, SNR increased when using a digital headstage compared to an analog headstage.

### System timing

When examining system latency, we found that the portable iBCI had increased variability in the timing of incoming spike data packets. An example of this intersample timing variability is shown in Figure 4. These delays are quantified across three testing days in Table 2.

**Table 2.** System delays (mean and standard deviation)

|  | **Standard (Lab)** | **Portable (Lab)** | **Portable (Home)** |
| --- | --- | --- | --- |
| **Intersample period (ms)** | 20.00 ± 0.29 | 22.06 ± 8.61 * | 20.00 ± 3.57 * |
| **SPD (ms)** | 5.83 ± 4.02 | 3.57 ± 3.45 * | 2.82 ± 1.79 *† |

*\* indicates significant difference from the standard iBCI. † indicates significant difference from portable iBCI tested in the laboratory.*

**Figure 4.**
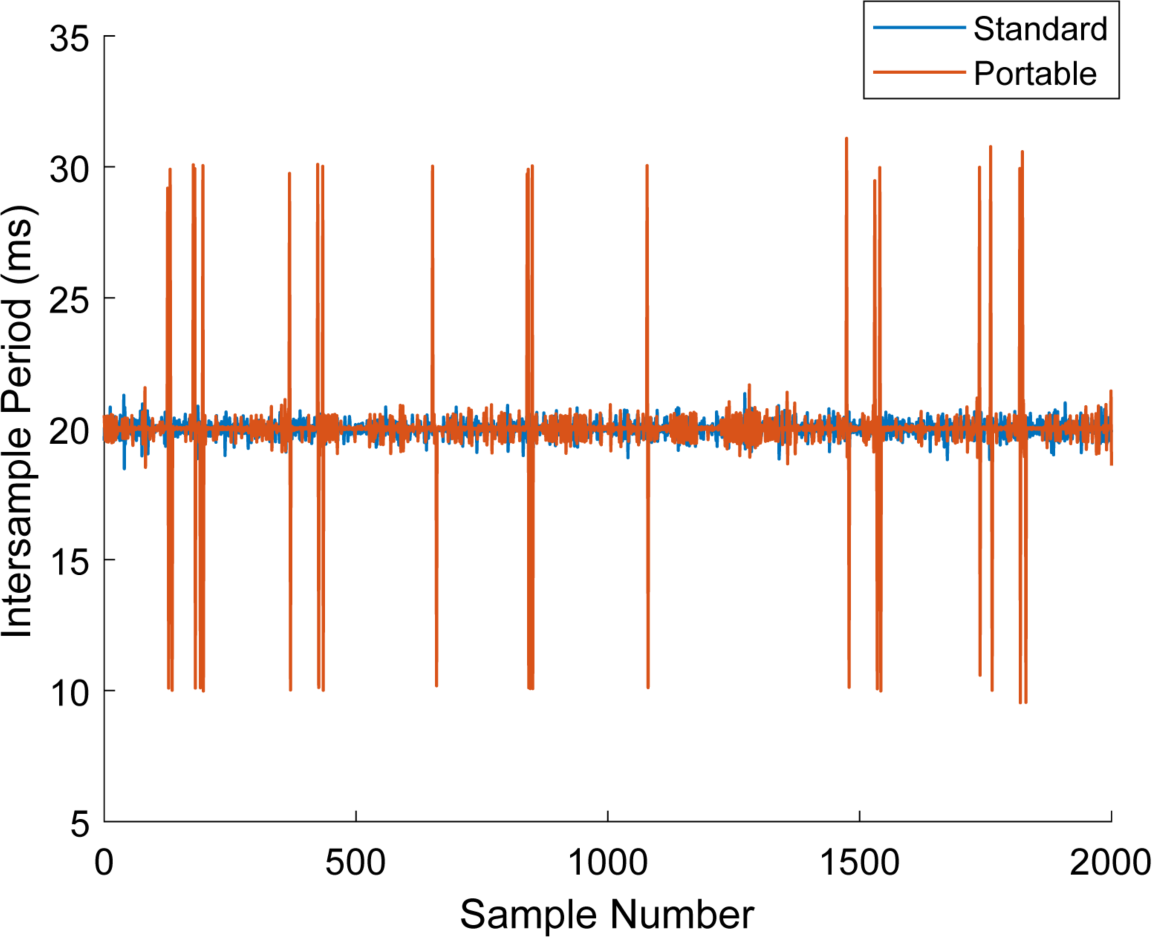
Intersample periods. Both the standard (blue) and portable (orange) iBCI systems are expected to output a binned spike count packet every 20 ms. Both systems exhibit a mean intersample period of approximately 20 ms in the representative timecourse of system timing, but the portable iBCI exhibits more variability with symmetric spikes at 10 and 30 ms indicating that packets are occasionally delayed, but not dropped.

The intersample periods between spike data packets were significantly different between recording systems (*F*(2, 98912) = 1586, *p* < 0.001), with post-hoc tests revealing less delay in the standard system as compared to the portable system at either the lab or home (both *p* < 0.001). Most of the difference between the three groups can be explained by the increased variance of the portable iBCI. While the mean intersample period was greatest during laboratory testing with the portable iBCI, most of this increased delay occurred during only one of the three test sessions. The mean intersample periods for each individual session were 25.7 ± 12.0 ms, 20.9 ± 7.3 ms and 20.0 ± 3.9 ms, respectively. The system processing delay (SPD) between spike count packets and subsequent decoded velocity commands was also significantly different across the three system conditions (*F*(2, 97357) = 7469, *p* < 0.001, all pairwise comparisons p < 0.001), with the standard system having the highest SPD and the portable system having the lowest SPD during in-home testing.

### Center-out task performance

The participant had considerable experience with the 2D center-out task prior to these sessions and in these experiments, success rates were generally unaffected by the type of system or testing location. The participant successfully completed 115/120 (95.8%) trials with the standard lab iBCI, 117/120 (97.5%) trials with the portable iBCI at the lab, and 114/120 (95.0%) trials with the portable iBCI at the participant’s home. Trajectories for all trials are shown in Figure 5. Sample performance can be seen in Supplementary Video 1.

**Figure 5.**
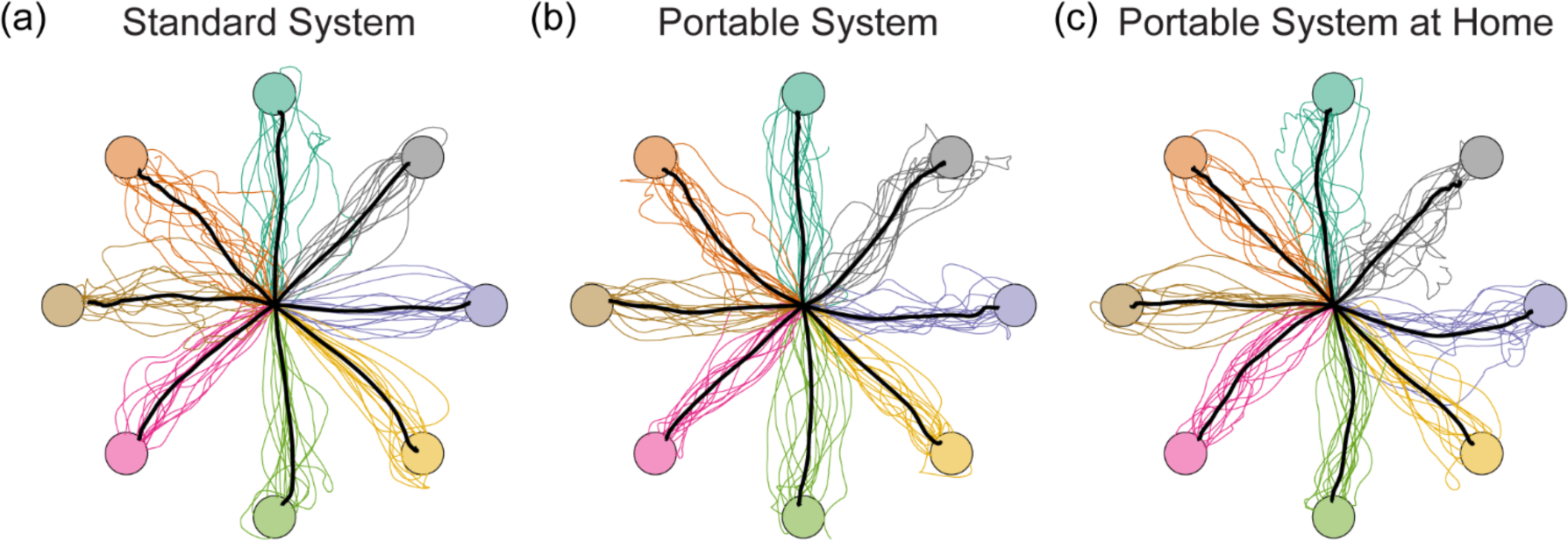
Center-out task trajectories from iBCI trials using (a) the standard laboratory system, (b) the portable system tested in the laboratory, and (c) the portable system tested at the participant’s home. Black lines indicate the mean trajectory across all trials to the respective target. Individual colored lines show trajectories from individual trials to targets with matching colors.

The mean and standard deviation values of time-to-target and normalized path length for successful trials and normalized mean decoded speed of all trials are summarized in Table 3. The time-to-target was significantly faster on the standard laboratory iBCI than the portable iBCI tested at the laboratory or the participant’s home (*F*(2, 343) = 12.3, *p* < 0.001, one-way ANOVA; post hoc *p* < 0.001 and *p* < 0.005 respectively). No significant difference was observed in the mean path lengths achieved with the three recording conditions (*F*(2, 343) = 0.363, *p* = 0.696). Normalized mean decoded cursor speed was significantly faster on the standard iBCI, as compared to the portable iBCI at the lab or the participant’s home (*F*(2,357) = 27.05, *p* < 0.001; post-hoc *p* < 0.001).

**Table 3.**
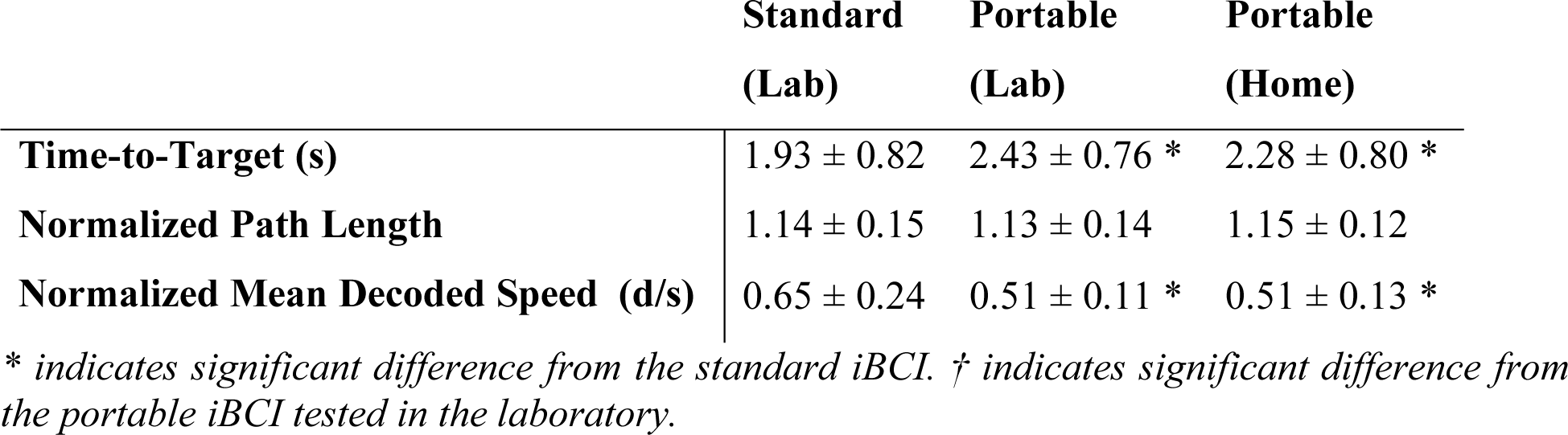
Time-to-target, normalized path length, and normalized trial mean decoded speed (mean and standard deviation)

### Functional evaluation

The participant was able to use the portable iBCI both in the lab and at his home for a variety of uses including computer painting, playing games, and typing. Examples of these tasks are shown in Figure 6. Figure 6a shows the participant using the system mounted to his wheelchair, while in Figure 6b it is positioned in his lap. Sample performance during iBCI painting and gaming can be seen in Supplementary Videos 2-3. The participant was also able to participate in a multi-player computer game with able-bodied competitors by connecting conventional USB gamepad controllers to the tablet using a USB hub.

**Figure 6.**
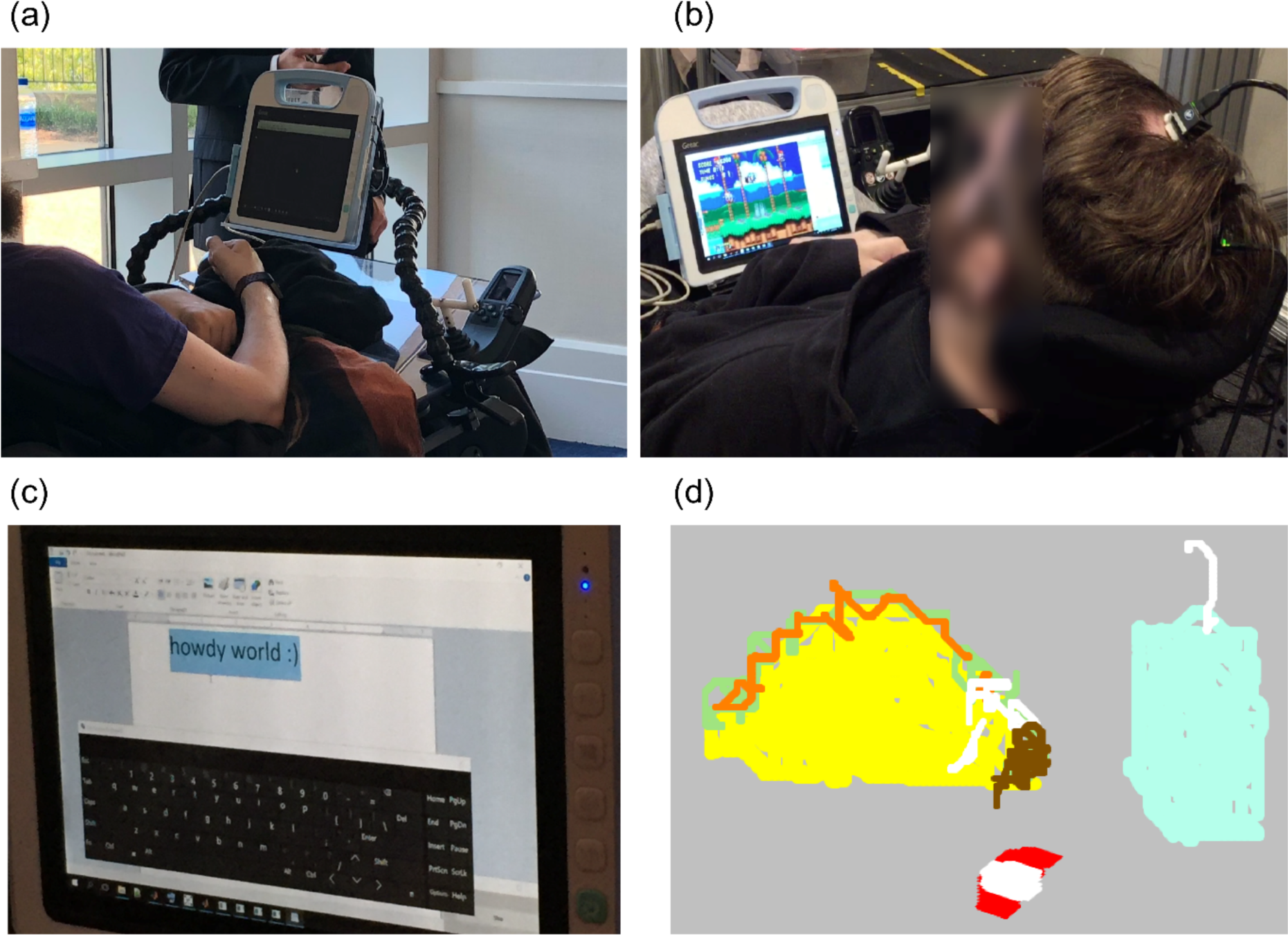
Functional use of the portable iBCI. (a) The participant performed the center-out task prior to freeform tasks. (b) The participant played computer games by converting decoded velocity commands to emulated keypresses. (c) The participant was able to type “howdy world:)” using an on-screen keyboard and iBCI mouse emulator. (d) The participant painted his favorite fast food taco order using an iBCI mouse emulator and a painting program.

The decoder calibration procedure alternated click and release targets across trials, resembling a click-and-drag behavior. This was well-suited for the painting task, but made it relatively difficult to click on buttons, such as to select different paint brushes and colors or during the typing task. Despite this difficulty, the participant was able to type the phrase “howdy world:)” using an on-screen keyboard, as seen in Figure 6c and successfully painted with a variety of colors, as seen in Figure 6d.

## Discussion

We demonstrated the feasibility of a portable iBCI that can fit on a user’s power wheelchair and be used for a variety of computer tasks such as typing, painting, and gaming. The system builds on FDA-cleared technology and includes only three major hardware components that are relatively lightweight and simple to setup: digital headstages, a signal processing hub, and a tablet PC. The small form factor and simplicity of the hardware could potentially pave the way for independent home iBCI use, or even mobile use of an iBCI in public settings.

The digital headstages used in the portable iBCI provided a small, yet statistically significant, increase in mean signal-to-noise ratio as compared to the analog patient cables and amplifiers traditionally used in laboratory iBCI systems. This increase in SNR can likely be explained by the observed decrease in baseline noise due to the fact that the digital headstages place the amplifier and digitizer at a much shorter distance to the electrodes, directly on top of the percutaneous electrode connectors, and without requiring cables between the percutaneous connector and amplifier. We unexpectedly found that the mean peak-to-peak voltage (*V*_pp_) was also reduced when using the digital headstages. This result can possibly be explained by the fact that the spike detection thresholds, which are calculated based on the signal *V*_RMS_, were lowered due to the reduced noise level, allowing signals with a smaller *V*_pp_ to be detected as spikes.

The participant in this study was able to complete a 2D cursor center-out task on the portable iBCI with a very high success rate indistinguishable from the success rate accomplished using the standard lab iBCI. Path lengths to each target were generally efficient and were also indistinguishable from those performed on the standard iBCI. However, trial completion times were significantly slower when using the portable iBCI. Several reasons may have contributed to this discrepancy in performance.

First, the portable iBCI system timing was more variable than the standard iBCI. The system was expected to output binned spike count data every 20 ms, but these data packets were sometimes delayed by 10 ms or more, an irregularity generally not seen on the standard iBCI. Despite this intersample period irregularity, the additional system processing and decoding delays within the iBCI software suite were actually more efficient on the portable iBCI, possibly due to the elimination of network transmission delays by consolidating the system onto a single computer. It is unclear if these timing irregularities affected the participant’s control strategy or introduced any feedback delays.

Second, in addition to the timing irregularities, the mean decoded speed was slightly slower when using the portable iBCI. One possible explanation for this reduction in decoded speed is the fact that the portable iBCI utilizes a tablet screen that is much smaller than the television display used with the standard system. Decoded velocity units were based on a fixed cursor coordinate system that visually scaled based on display size. However, the participant observed the cursor move in a physically smaller workspace on the portable iBCI and may have imagined slower velocities as a result. However, these slower velocities did not impede the participant’s ability to successfully complete trials and he was able to demonstrate a variety of functional tasks with the portable iBCI, both in the lab and at the home, including drawing, gaming, and typing.

There are several limitations of the portable iBCI that could be improved or eliminated in future iterations of the system. First, while we were able to test the iBCI both in the lab and in the participant’s home, as per our trial protocol, all tests required an experimenter to be present to set up, configure, and monitor the system. The software interface is adapted from the standard system, which was designed for the flexibility necessary to run a wide variety of experiments. This resulted in a complicated interface intended to be run by an experimenter with many unnecessary features for an end-user. Further iterations of the iBCI software suite are required to shift to a user-centric design and allow for independent use of the system by iBCI users and caregivers.

Second, we found that the neural data was received by the iBCI software suite with irregular timing that exhibited more variability than the standard iBCI. The standard iBCI includes dedicated neural signal processors (NSPs) that run a real-time operating system. The NSPs receive continuously sampled neural data from the amplifiers through a fiber optic connection and output extracted spike snippets in real-time to a networked PC over ethernet. Additionally, the standard iBCI software suite is distributed across several high-performance PCs. This is contrasted with the portable iBCI, in which all neural signal processing and iBCI software runs on the Windows 10 tablet PC. Continuously-sampled neural data is transmitted to the tablet from the wNSP hub over USB 2.0. The USB 2.0 specification defines a maximum signaling rate of 480 Mb/s [26]. The portable system streams 16-bit neural data sampled at 30 kHz from up to 256 electrode channels to the tablet over USB, which requires a minimum bandwidth of 123 Mb/s, or 26% of the maximum signaling rate permitted by USB 2.0. While this is theoretically enough bandwidth, a faster protocol such as USB 3.x or gigabit ethernet would enable expansion to higher channels counts.

Additionally, the tablet PC has limited resources to receive and process the neural data, run the iBCI software suite to decode neural spikes and generate velocity commands, and run external programs that the user wishes to control. Running these tasks on relatively low-performance tablet hardware may result in poor performance, manifested by irregular timing and poor battery life. Additional processing power may be needed to study higher degree-of-freedom control or to interface with external devices. There are several potential options that may improve performance. The most straightforward option is to use a more powerful tablet PC. The tablet in this study was chosen primarily because it had already undergone IEC 60601-1 testing and is considered medical-grade. The wNSP hub is designed to electrically isolate the tablet from the digital headstages, and thus the requirement for an IEC 606061-1 compliant tablet can potentially be relaxed though this would need to be confirmed through electrical safety testing of a complete system. A tablet with a faster processor would likely improve performance. Additionally, while it would not be desirable to add additional components to the system, it may be possible to offload some of the neural signal processing and decoding to a small secondary microcontroller networked with the tablet PC. This would free up resources on the tablet PC for applications that the user directly interacts with. Finally, in a home environment, it may be preferable for the iBCI to network with the user’s personal computer. In this scenario, the tablet PC would perform neural signal processing and decoding, but would send commands such as emulated mouse movements and clicks to an external computer running the user’s choice of software.

The functional demonstrations presented here suggest that an iBCI user could use the portable iBCI for a variety of independent tasks, including communication tasks, creative tasks such as drawing, and recreational tasks such as gaming and web browsing. It is worth noting that the focus of this study was in the development and evaluation of portable iBCI hardware rather than algorithm development for optimal computer access. Therefore, this work is complementary to that of other research groups that have demonstrated high levels of performance for computer cursor control and communication (e.g. [1], [2]) including integration of laboratory hardware systems with commercially available tablets through wireless communication [19]. In future work, we would like to investigate the portable iBCI in a take-home study to evaluate independent use of the system by the iBCI user outside of a controlled setting, which may include use at home or mobile use in public settings.

## Data Availability

Data supporting these findings as well as software routines to analyze these data are available from the corresponding author upon reasonable request.

## Acknowledgements

We would like to thank our study participant for making this work possible. The authors also thank to RNEL neurosurgeon Elizabeth Tyler-Kabara and lab members Kristin Quick, Angelica Herrera, and Chris Hughes for their contributions to electrode array implantation and data collection, as well as Gina McKernan for discussions about statistical analyses. We would like to thank Blackrock Microsystems employees Charles Dryden and Hyrum Sessions for contributions to hardware and firmware development.

## Disclosure of interest

Robert Franklin is an employee of Blackrock Microsystems LLC, a company that may be affected by the research reported in the enclosed paper.

## Funding

This work was supported by the Defense Advanced Research Projects Agency (DARPA) and Space and Naval Warfare Systems Center Pacific (SSC Pacific) under Contract N66001-16-C-4051. Any opinions, findings and conclusions or recommendations expressed in this material are those of the authors and do not necessarily reflect the views of DARPA or SSC Pacific.

## Notes

### Clinical Trial

NCT01894802

### Author Declarations

All relevant ethical guidelines have been followed and any necessary IRB and/or ethics committee approvals have been obtained.

Any clinical trials involved have been registered with an ICMJE-approved registry such as ClinicalTrials.gov and the trial ID is included in the manuscript.

